# Association between temperature and back pain among lumbar disc herniation patients

**DOI:** 10.1101/2020.01.15.20017558

**Authors:** Cai Chen, Dandan Wang, Fanjie Liu, Hao Qin, Xiyuan Li, Fan Bu, Wei Li, Bin Shi, Shengnan Cao, Jianpeng An, Xiangwei Meng, Zhixiang Ma, Leilei Dong, Qinghao Zhang, Tao Wang

**Author notes:** Corresponding author: Wei Li: Biomedical Engineering Institute, School of Control Science and Engineering, Shandong University, Jinan 250061, China, Bin Shi: Bone Biomechanics Engineering Laboratory of Shandong Province, Neck-Shoulder and Lumbocrural Pain Hospital of Shandong First Medical University, Shandong Medicinal Biotechnology Center, Shandong First Medical University & Shandong Academy of Medical Sciences, Jingshi Road 18877, Jinan 250062, Shandong Province, China.

## Abstract

**Purpose:** This paper was designed to critically provide empirical evidence for the relationship between temperature and intensity of back pain among people with lumbar disc herniation (LDH).

**Methods:** Distributed lag linear and non-linear models (DLNM) was used to evaluate the relationship between lag-response and exposure to ambient temperature. Stratification was based on age and gender.

**Results:** When daily average temperature was on the rage of 15-23□, the risk of hospitalization was at the lowest level for men group. About below 10□, risk for male hospitalization could keep increase when lag day were during lag0-lag5 and lag20-lag28. 40<age≤50 group was little affected when they exposed to ambient temperature.

## Introduction

Low back pain (LBP) is one of the common diseases that affect severely people’s health, which tends to occur among middle-aged and elderly people[1, 2]. The prevalence of LBP increased with age, but remained unchanged at a certain age. The prevalence of LBP is increasing at an alarming rate in all age groups. It is reported that about 15%∼ 20% of adults had experienced back pain during one single year and 50% to 80% experience at least one episode of back pain during their lifetime[3]. Survey conducted in a university in the south of China has shown that about 731/3770 (19.39%) of all participants had chronic low back pain and more female students had chronic low back pain[4]. LBP is increasingly recognized as a serious, worldwide public health concern. One published research showed that back pain experienced by patients were the one of strongest factors associated with this insomnia, which seriously affected patient’s quality of sleep[5]. In addition, there is accompanied with some neurological symptoms among LBP patients, such as radicular pain, radiculopathy and lumbar spinal stenosis[6, 7]. LBP has become one of the leading causes of years lived with disability in most countries and age groups[6]. It was predicted that low back pain prevalence and disability increased markedly over the past 25 years and would likely increase further with population aging[8].

Thus, determining the factors that could trigger the pain intensity are particularly crucial. It was suggested that there was an association between ambient temperature and the sensitivity and intensity of pain[9-11]. However, on the contrary, some previous research found that temperature didn’t influence the pain’s intensity and sensitivity[12-14]. Therefore, this primary purpose of this paper was designed to critically provide empirical evidence for the relationship between temperature and intensity of back pain among people with lumbar disc herniation (LDH).

## Method

Data about patient’s age, gender, diagnostic logout, admission time, discharge time, residence area and work area (residence area and work area were used to ensure research area) from 2016 to 2019, obtained from Neck-Shoulder and Lumbocrural Pain Hospital. Temperature data at the same period was attained from Shandong Environmental Protection Department. In our study, people with low back pain who were admitted to hospital and developed the lumbar disc herniation, were brought into this study, and were seen that the intensity of pain were acutely exacerbated.

Distributed lag linear and non-linear models (DLNM) describes the associations showing potentially non-linear and delayed effects in time series data, which has been widely used to evaluate the relationship between lag-response and exposure to ambient temperature[15-18]. DLNM was used to analyze the association between temperature and its lag-response for LDH patient’s with back pain. The DLNM was presented as follow:

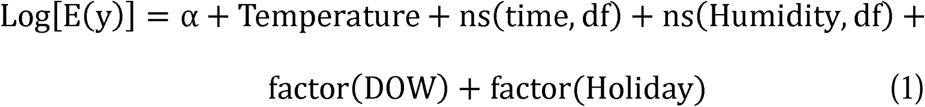

Where Log[.] means connected function, E(y) is the expected people who visited hospital, α is the constant term, Temperature represents the daily average temperature, time is the time variable, ns(.) represents natural cubic splines, df means the freedom of degree. DOW is the day of week, and Holiday is holiday variable. According to previous research, there was the lag effect of temperature on people’s health, therefore, lag effect of daily average temperature was considered in this study[18, 19]. Stratification based on age and gender was also considered. All the statistical analyses above mentioned were finished in *Rstudio* software.

## Result

### Descriptive result

**Table 1** describes the quartile of daily average temperature and relative humidity. The maximum and minimum of daily average temperature in Jinan during 2017-2019 were 33.4□ and −9.1□, respectively. The maximum and minimum of relative humidity during research period were 14 and 98, respectively. **Figure 1** presents the three-dimensional percentage bar chart based on age and gender. It can be found that in age≤30 group and 30<age≤40 group, the amount of men was more than that in women, while it was on the contrary among 40<age≤50, 50<age≤60, 60<age≤70 and age>70 group. **Figure 2** and **Figure 3** present the proportion of unhealthy habits and chronic diseases.

**Table 1.** Quartile of daily average temperature and relative humidity

|  | Min | P<sub>25</sub> | P<sub>50</sub> | P<sub>75</sub> | Max |
| --- | --- | --- | --- | --- | --- |
| Temperature | -9.1 | 6 | 16.5 | 25.3 | 33.4 |
| Humidity | 14 | 36 | 48 | 64.75 | 98 |

Min: minimum; Max: maximum; P25: the first quartile; P50: the second quartile; P75: the third quartile;

**Figure 1.**
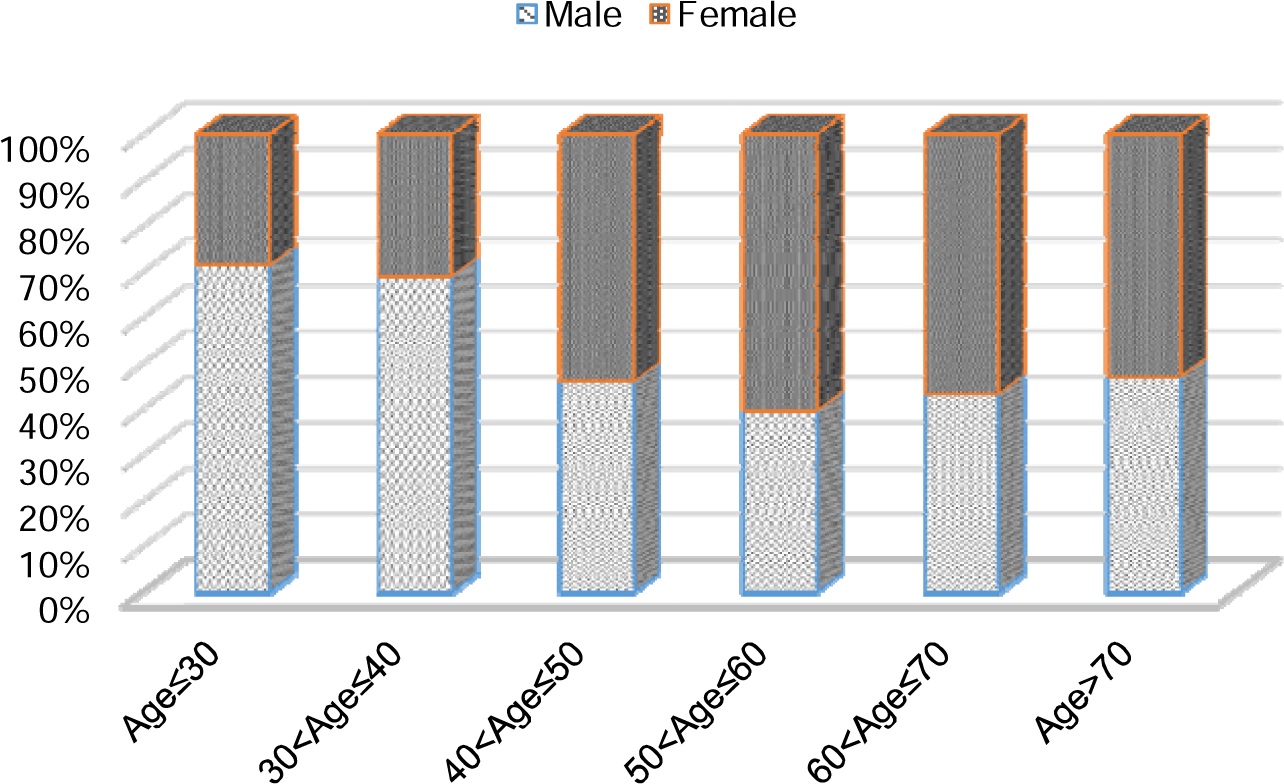
Three-dimensional percentage bar chart of age and gender

**Figure 2.**
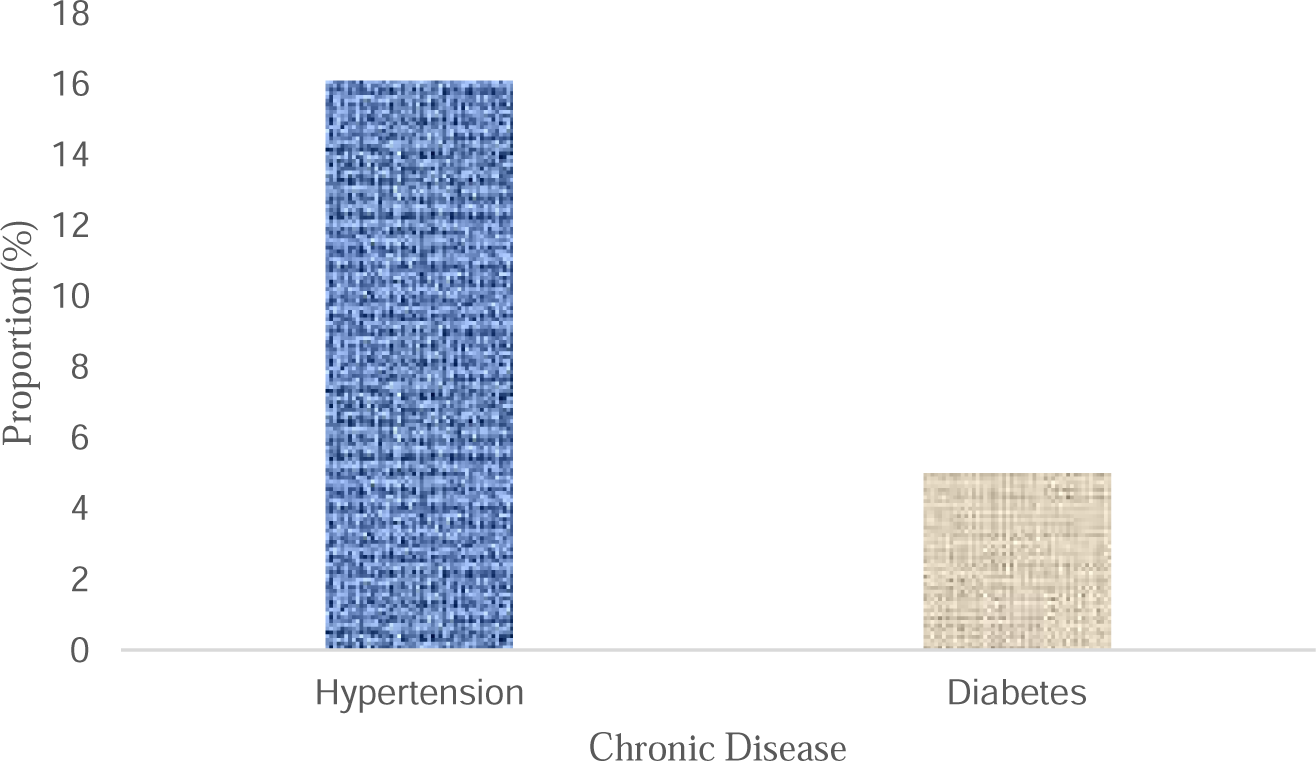
The proportion of chronic disease among whole amount

**Figure 3.**
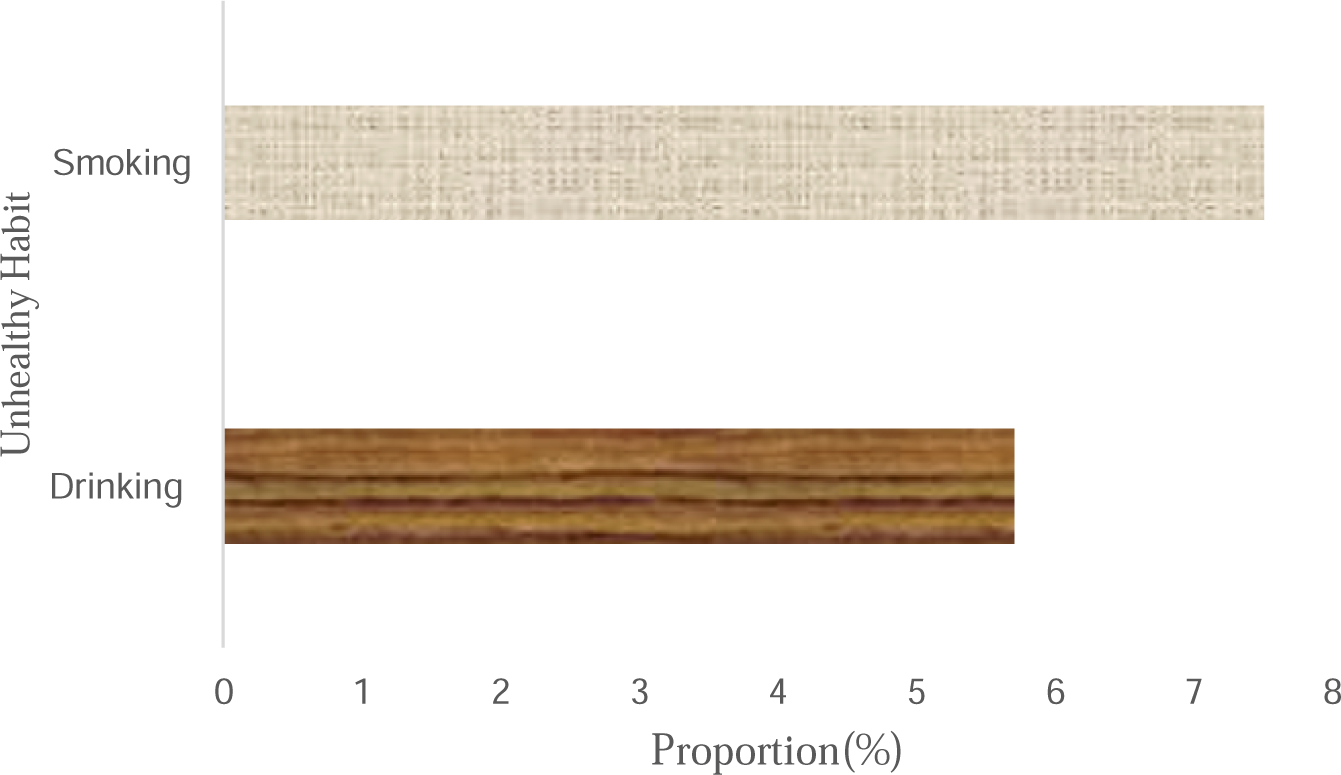
The proportion of unhealthy habits among whole amount

### DLNM Result

The results of lag-response curve of temperature were set out in **Figure 4**. As can be seen clearly from the Figure 4, response to exposure to about 25-30□ was different between men group and women group. **Figure 5** illustrates the effect of temperature on the hospitalization for LDH patients with back pain. When daily average temperature was on the rage of 15-23□, the risk of hospitalization was at the lowest level for men group, while for women group, temperature was above 15□, the risk was gradually decreased. From the contour plot of lag-response to temperature, when temperature was about below 10□, risk for male hospitalization could keep increase when lag day were during lag0-lag5 and lag20-lag28, similarly, temperature was about over 21□, lag effect would exacerbate the risk of hospitalization (**Figure 6**).

**Figure 4.**
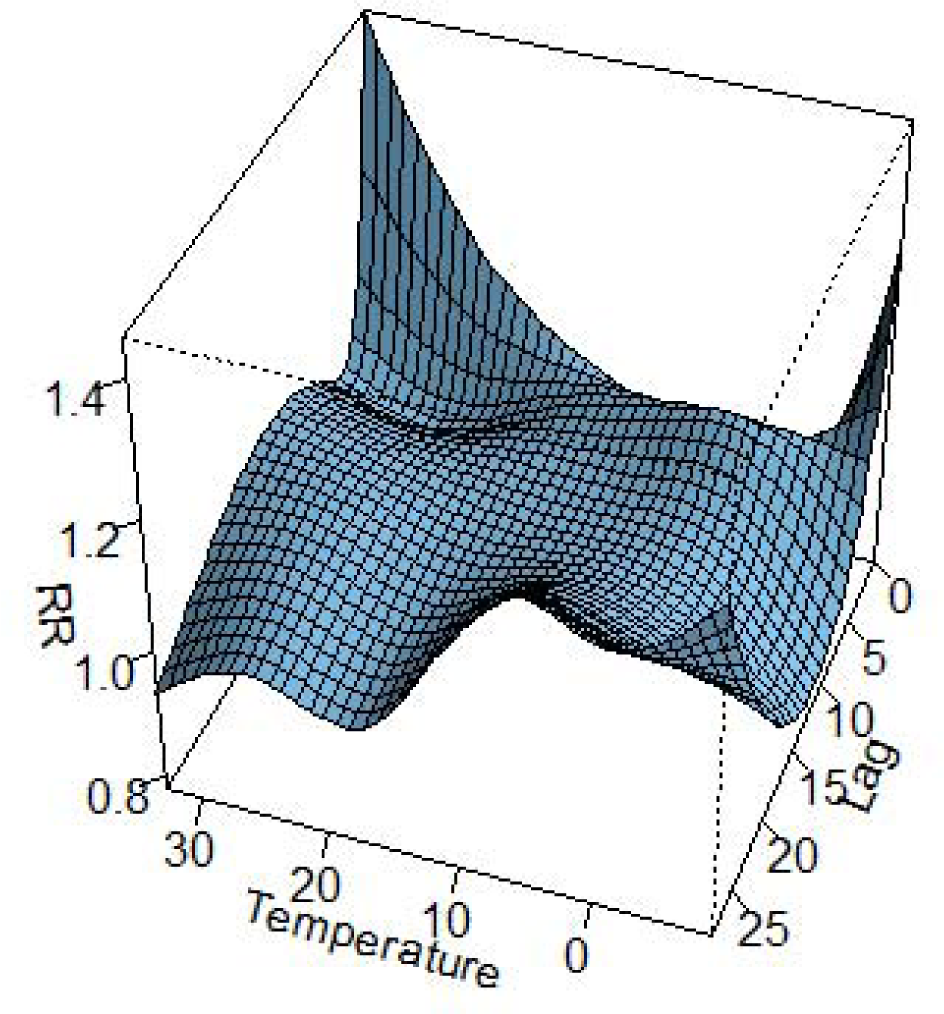
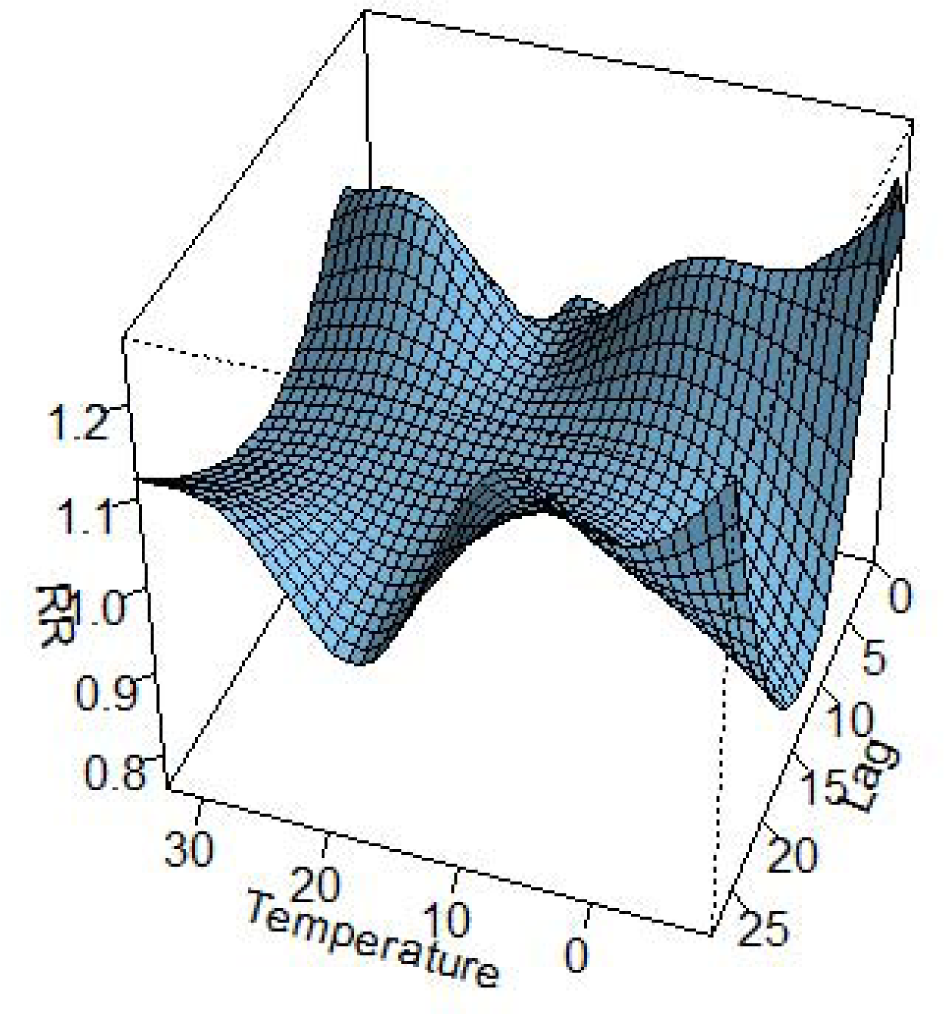
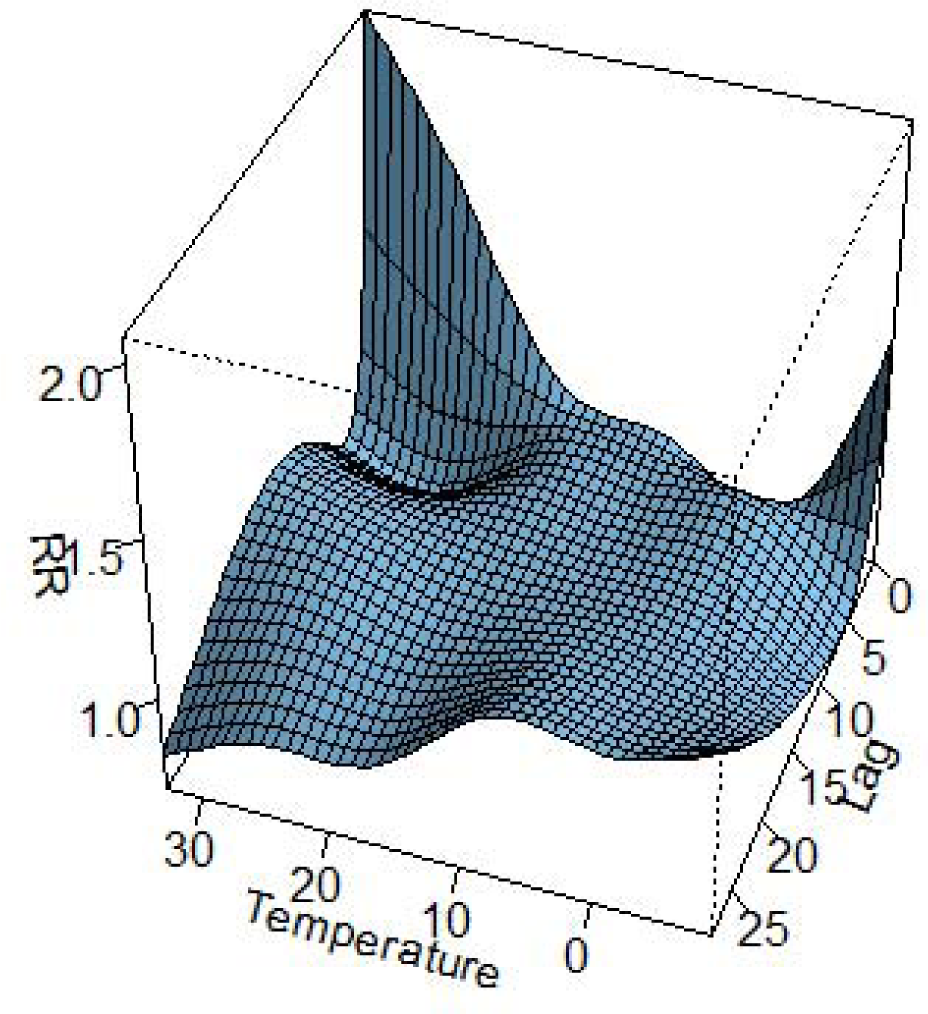
Three-dimensional lag-response curve of temperature (A: whole; B: Men; C: Women) *RR: relative risk;*

**Figure 5.**
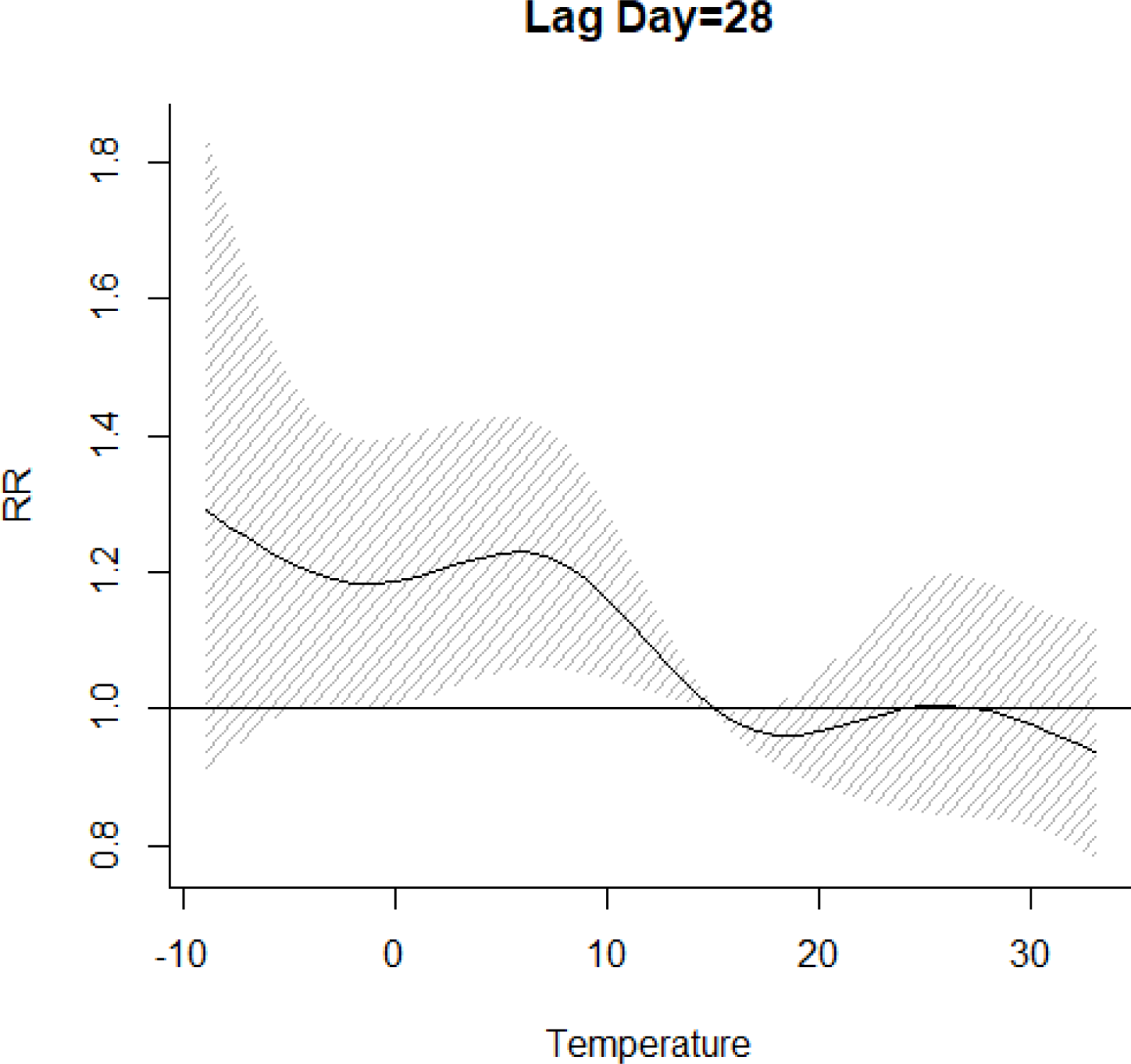
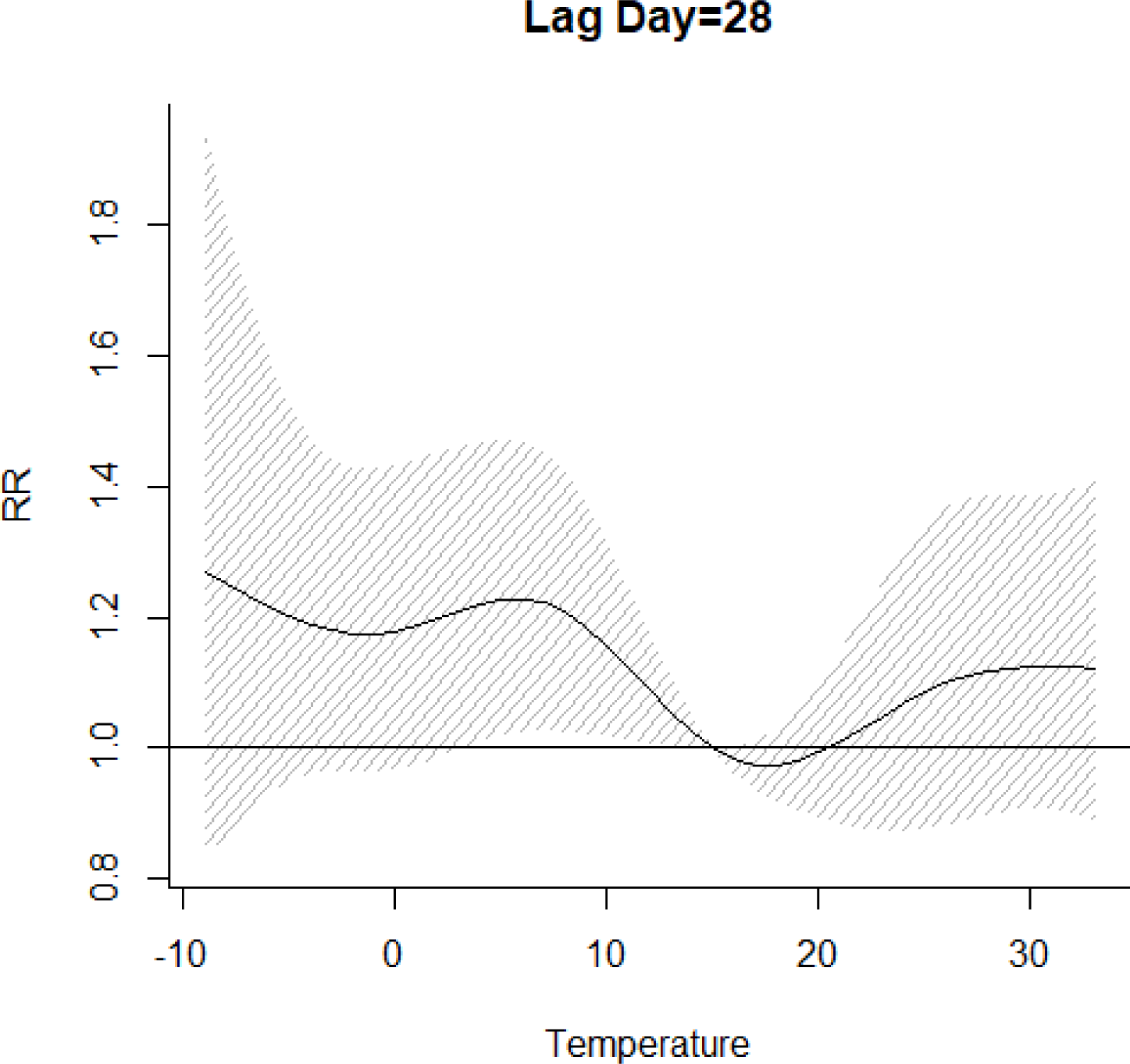
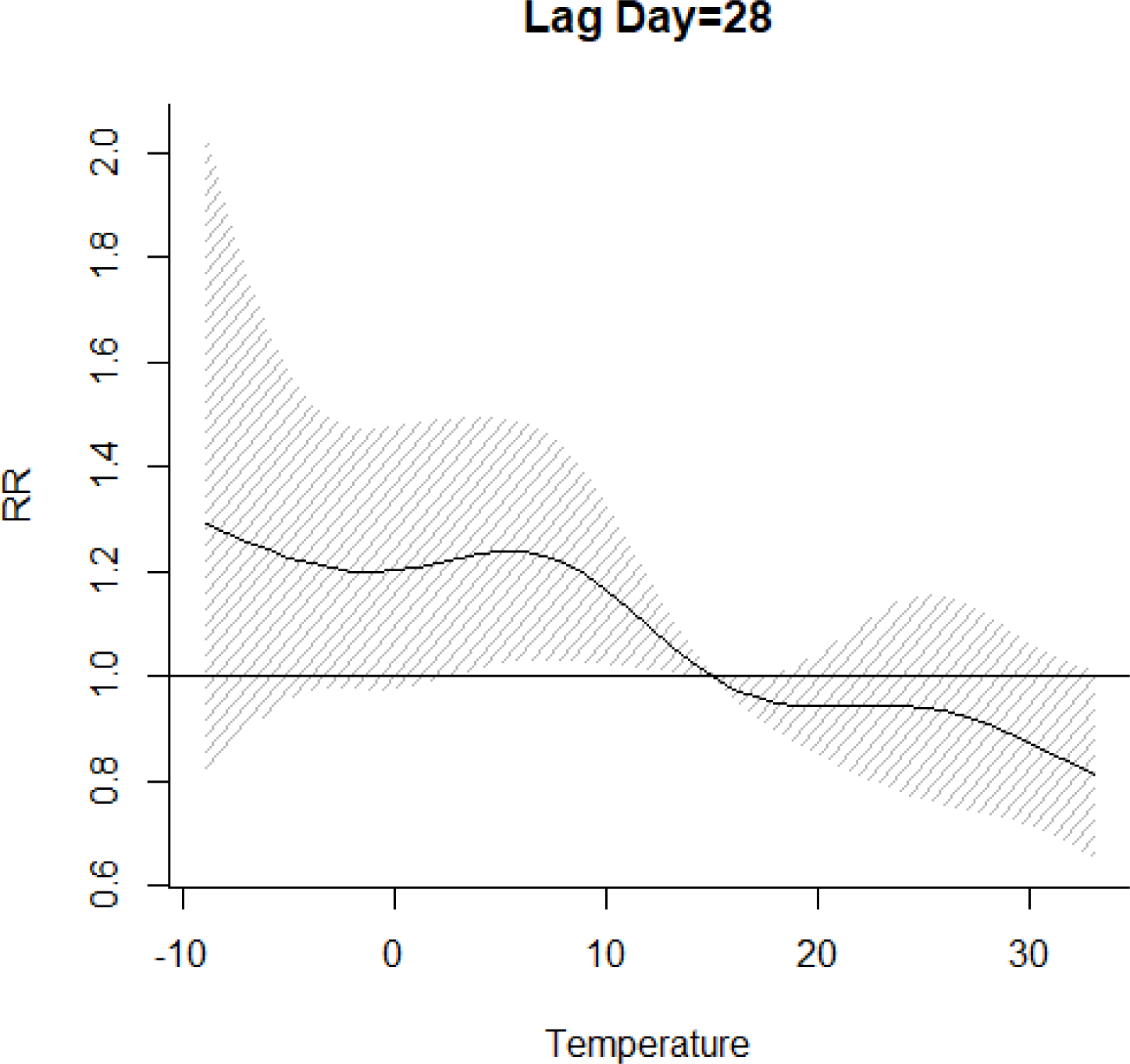
The effect of temperature on the hospitalization when lag day was on 28 (A: whole; B: Men; C: Women) *RR: relative risk*

**Figure 6.**
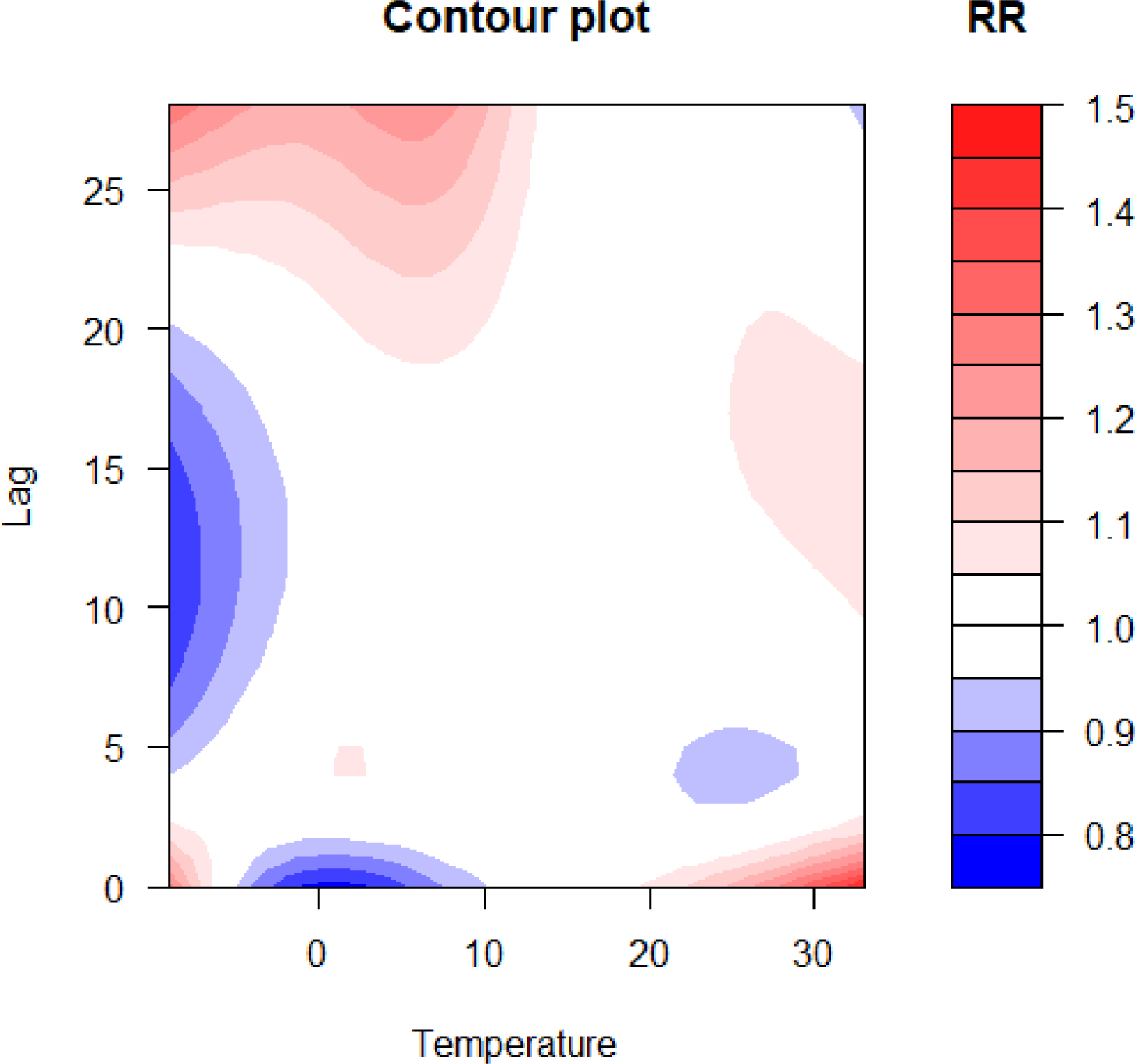
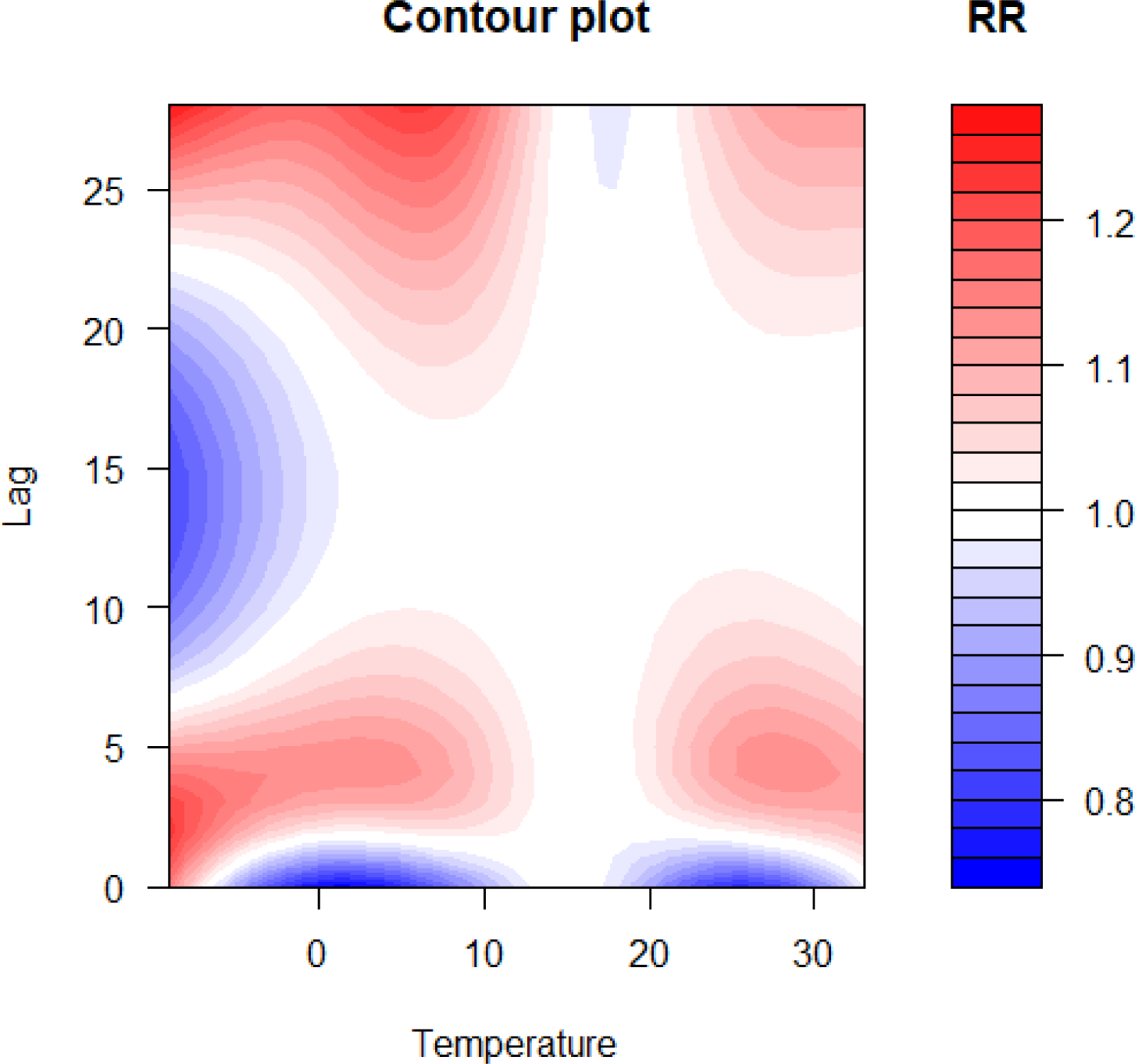
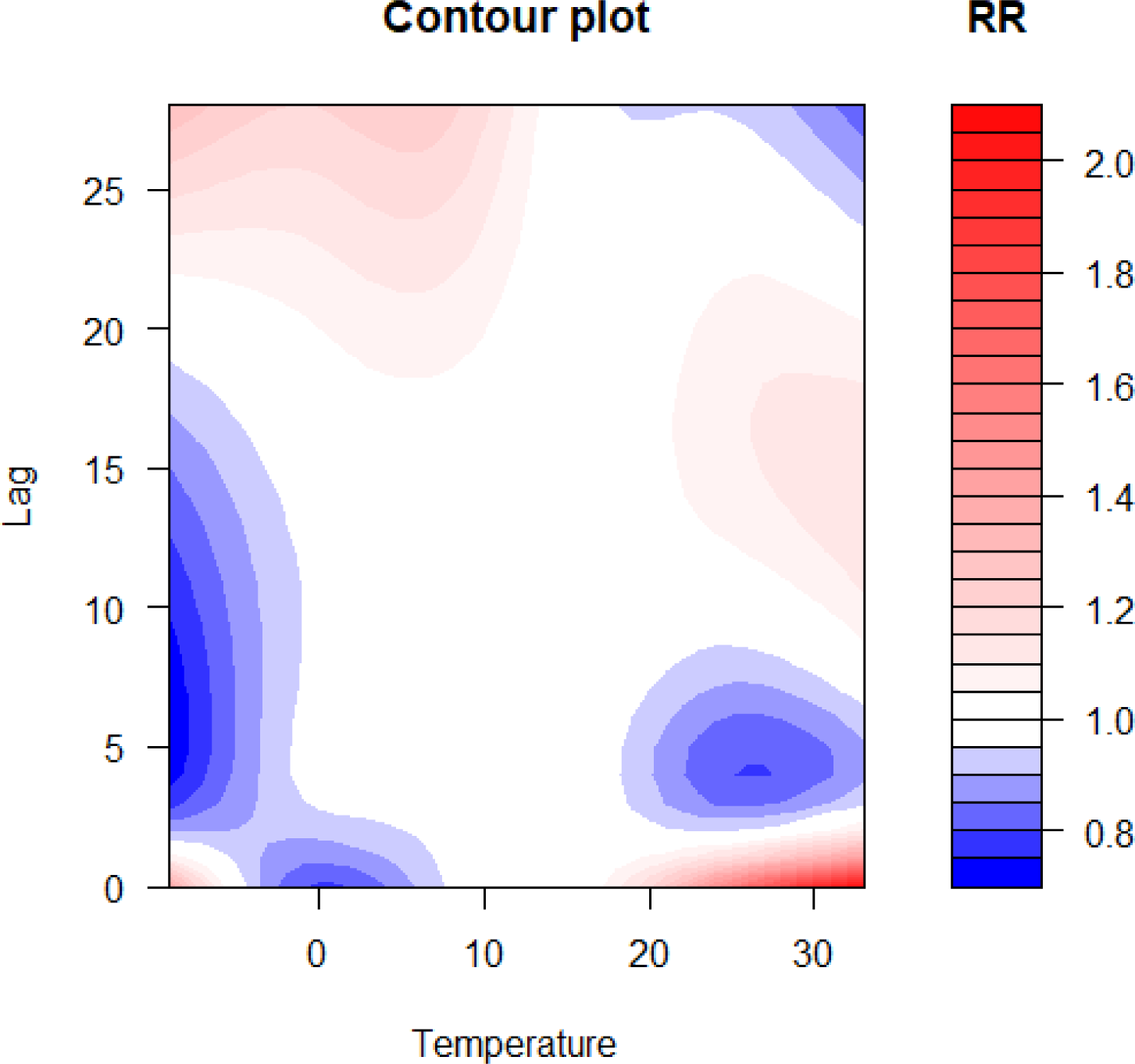
Contour plot of lag-response about temperature on hospitalization (A: whole; B: men; C: women) *RR: relative risk*

The three-dimension plots of lag-response to temperature based on stratification of age can be compared in **Figure 7**. Contour plots of lag-response to temperature based on the age stratification were displayed in **Figure 8**. What is striking about the Figure 8 is that 40<age≤50 group was little affected when they exposed to ambient temperature. The effect of temperature on the hospitalization when lag day was on 28. The effect of temperature on the hospitalization when lag day was on 28 was displayed on **Figure 9**.

**Figure 7.**
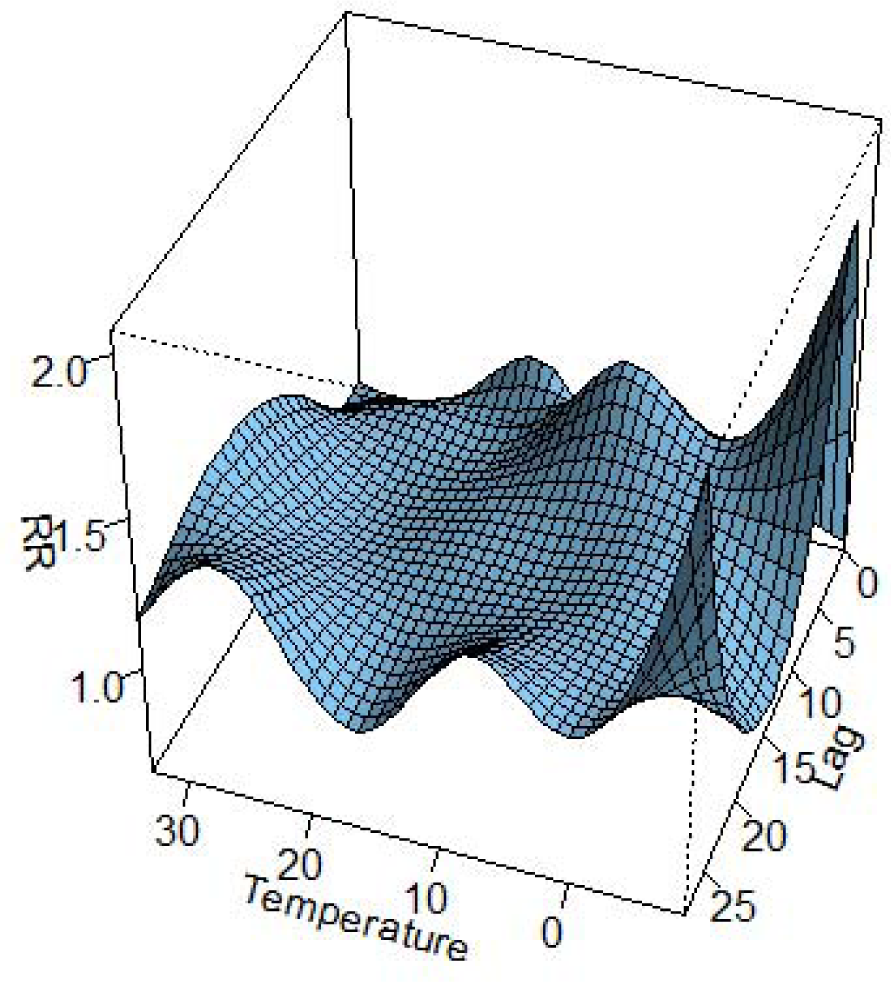
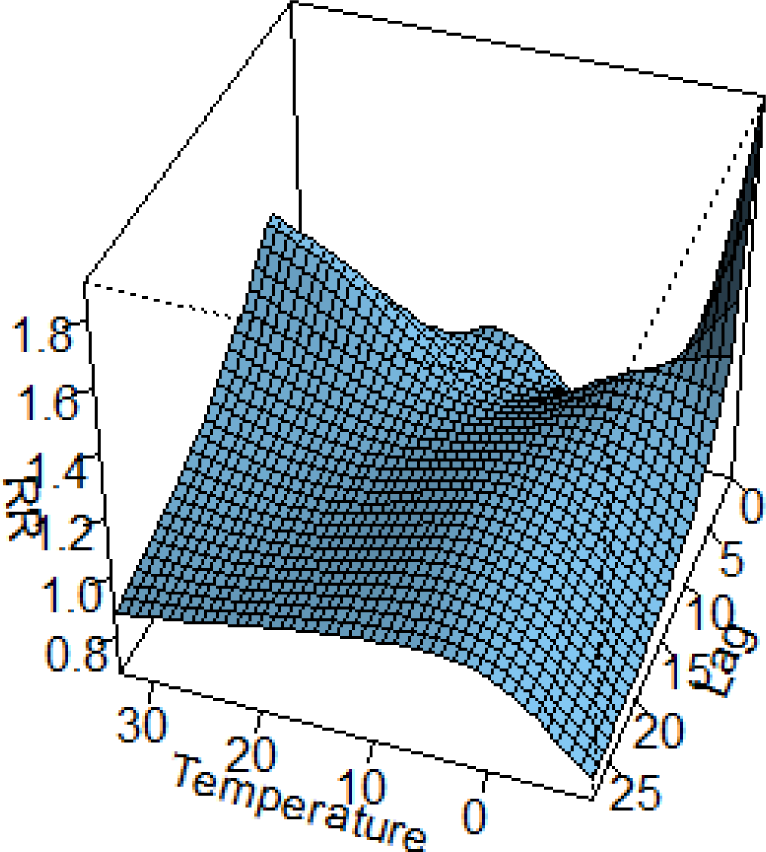
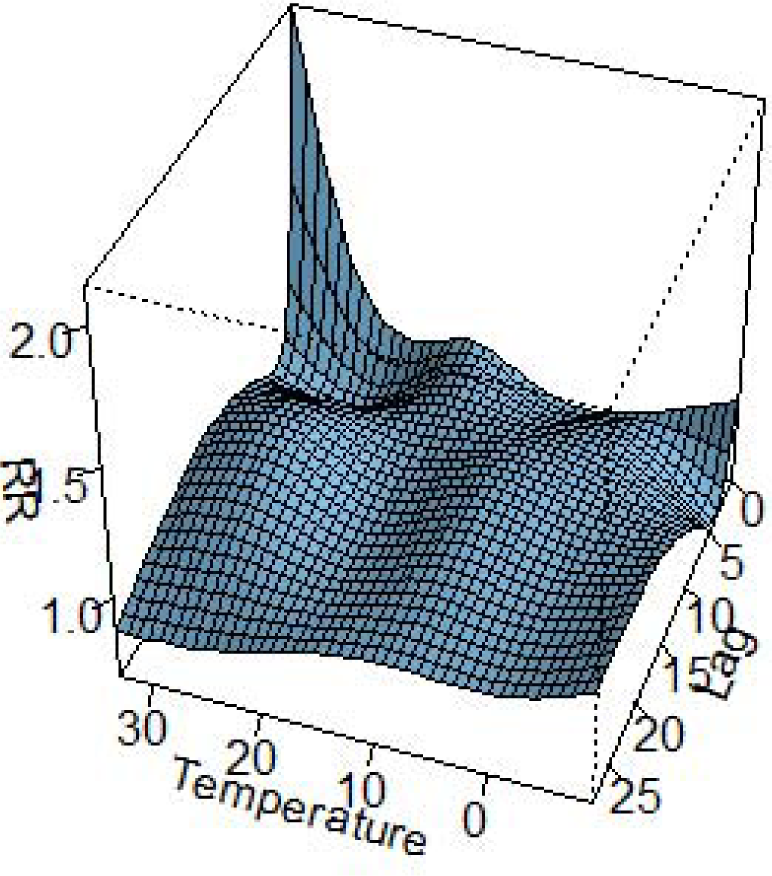
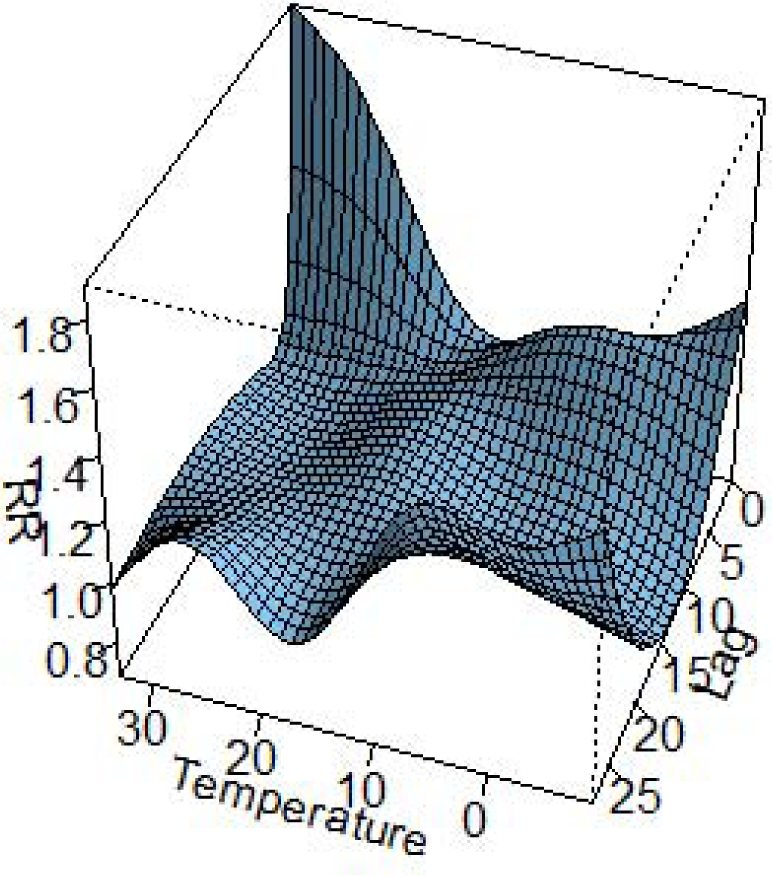
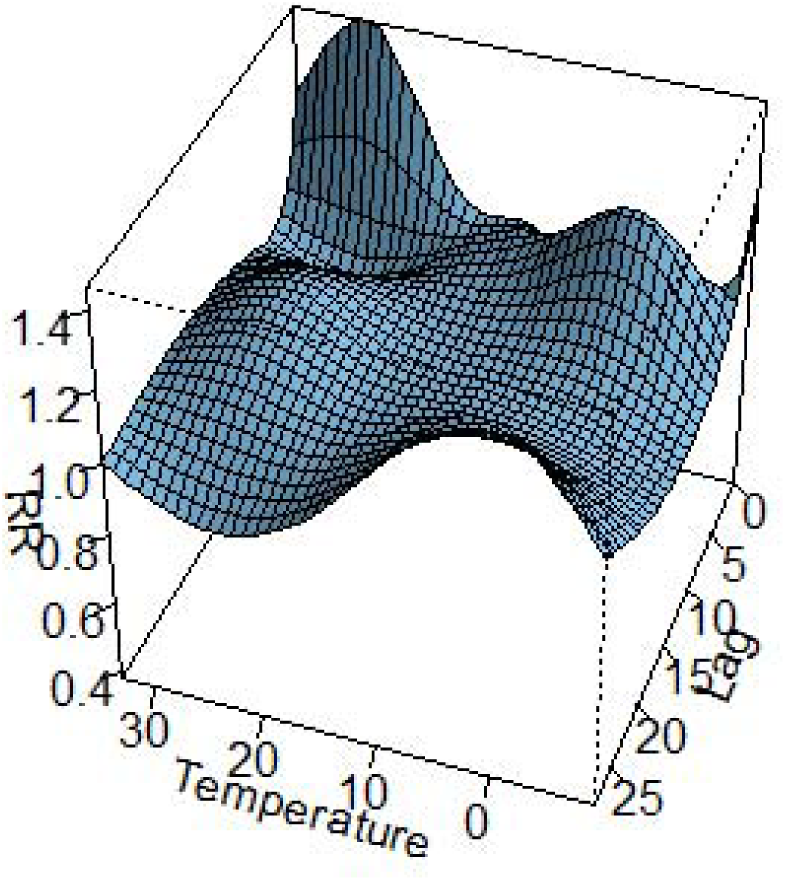
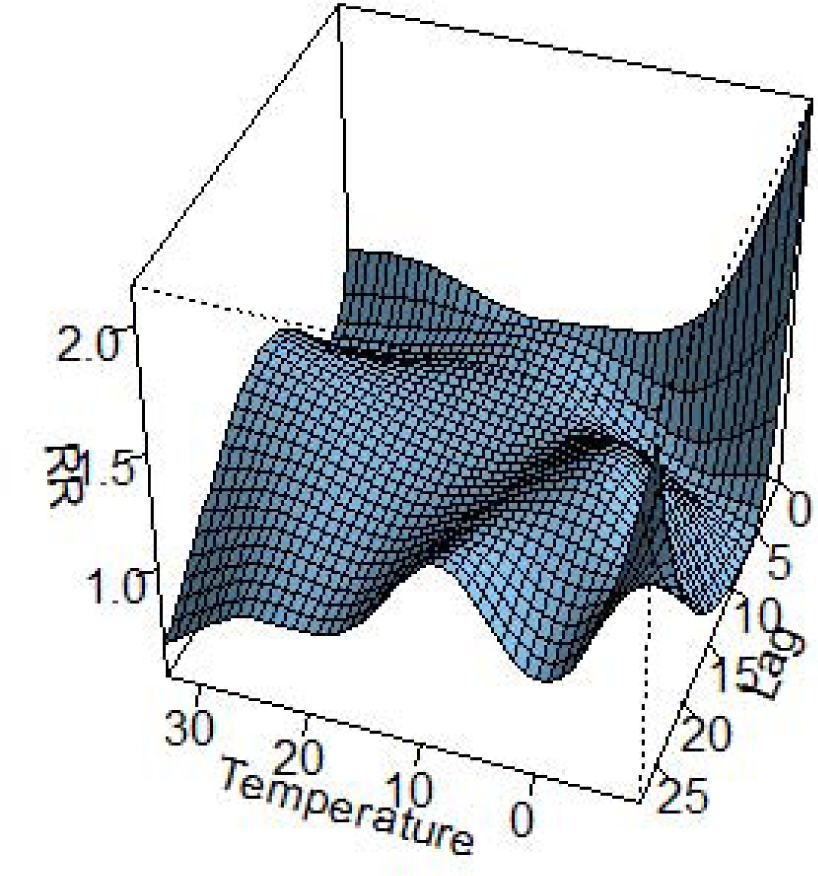
The three-dimension of lag-response to temperature based on stratification of age (A: age≤30 group; B: 30<age≤40 group; C:40<age≤50; D:50<age≤60; E: 60<age≤70; F: age>70 group)

**Figure 8.**
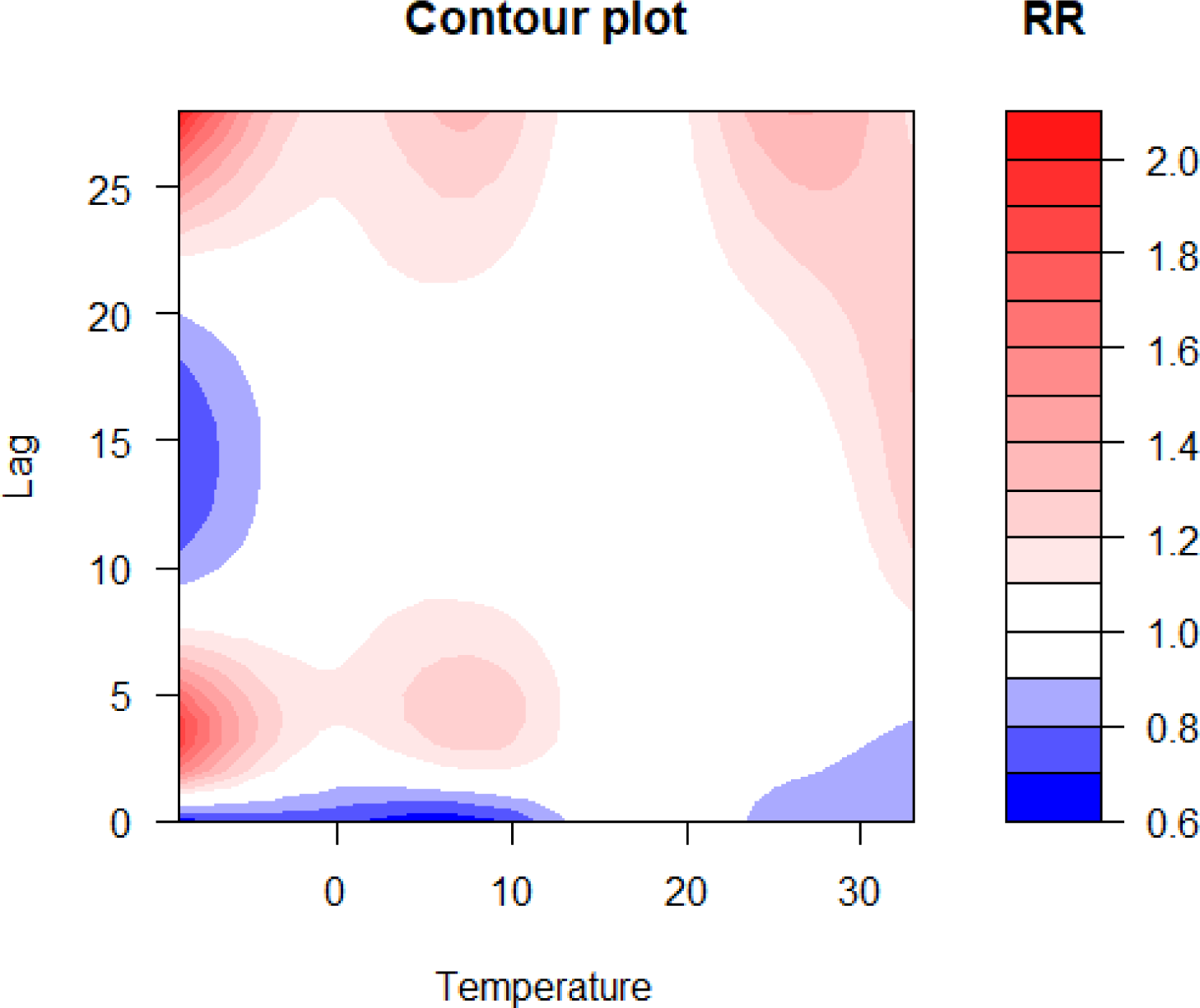
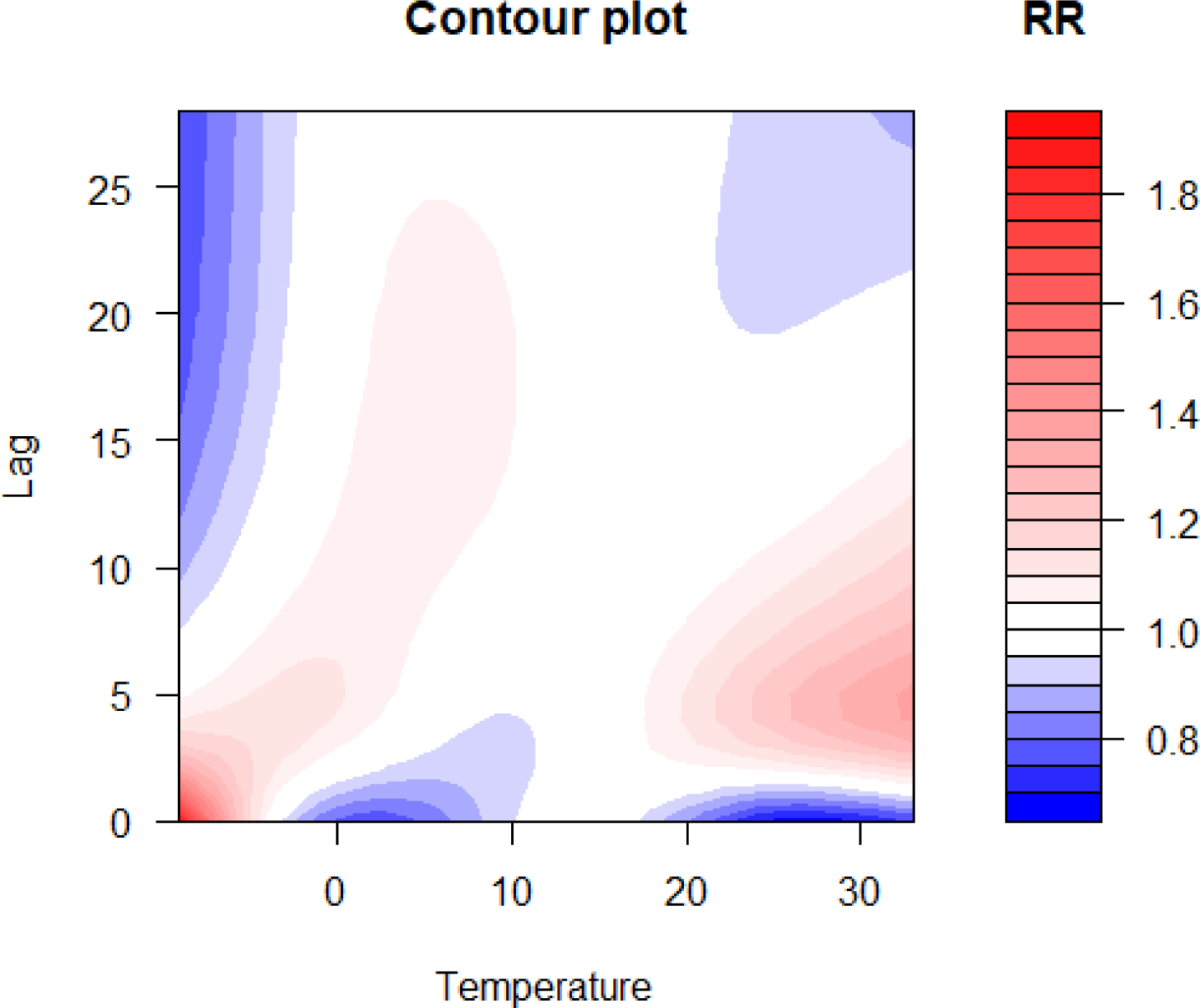
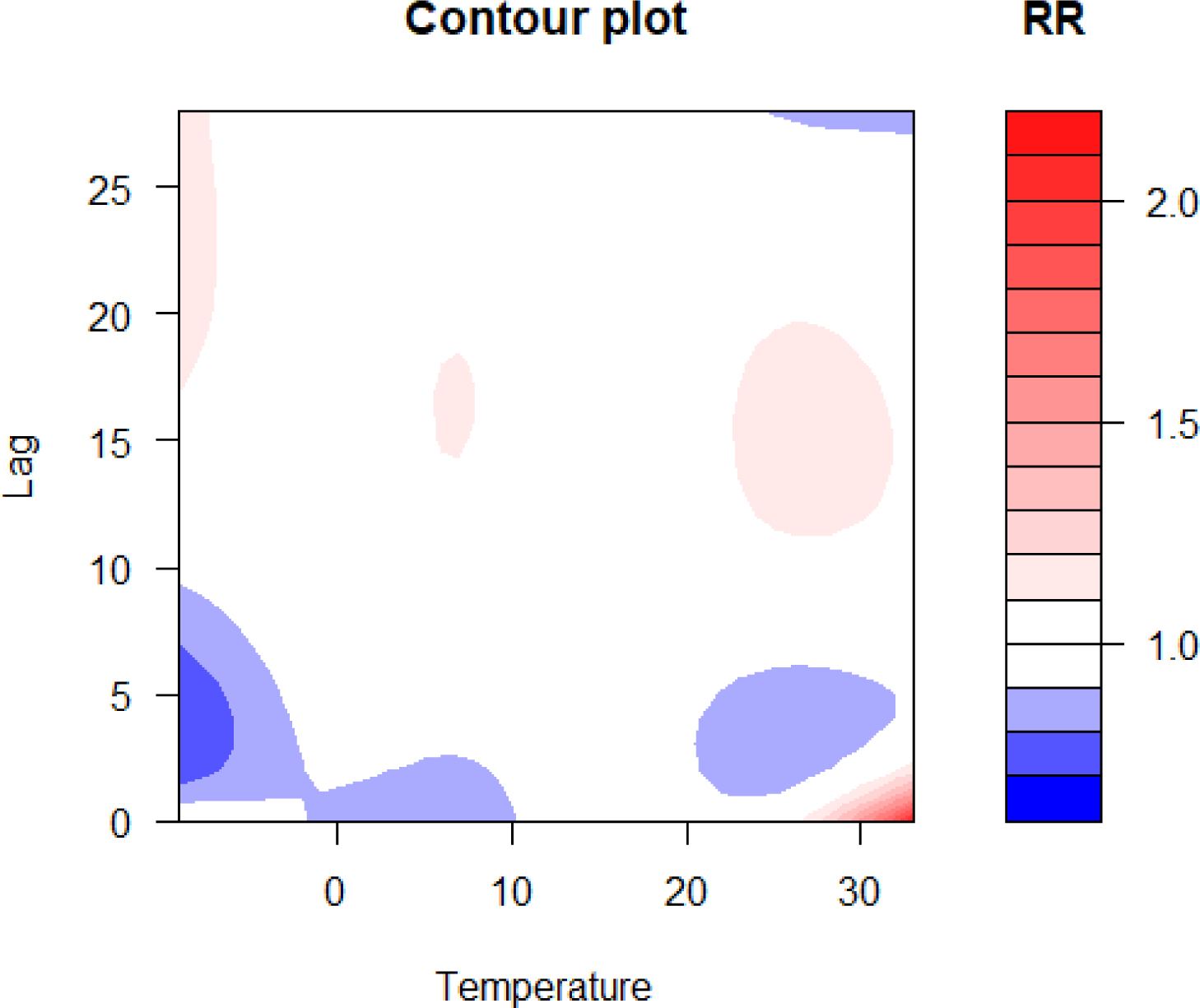
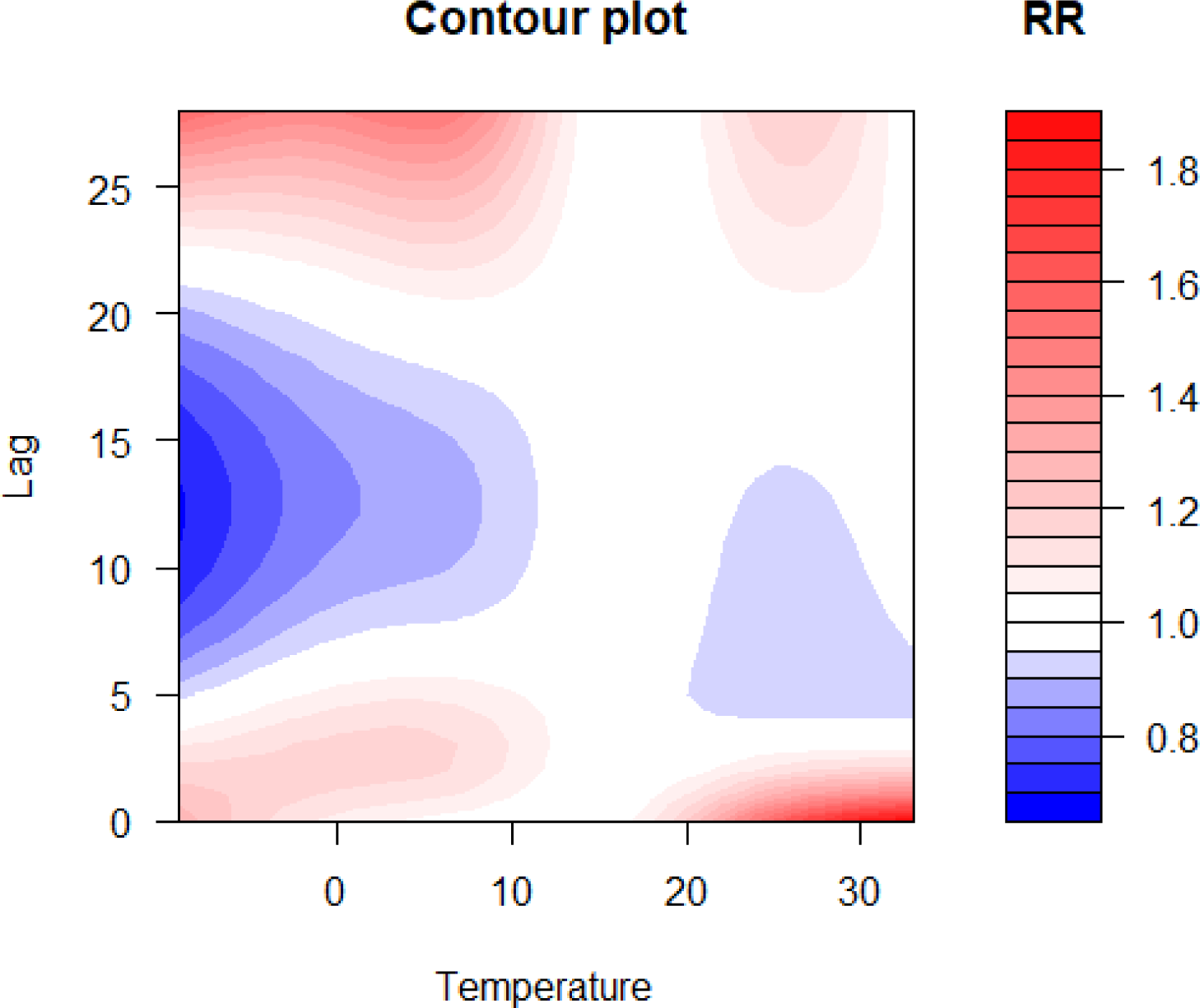
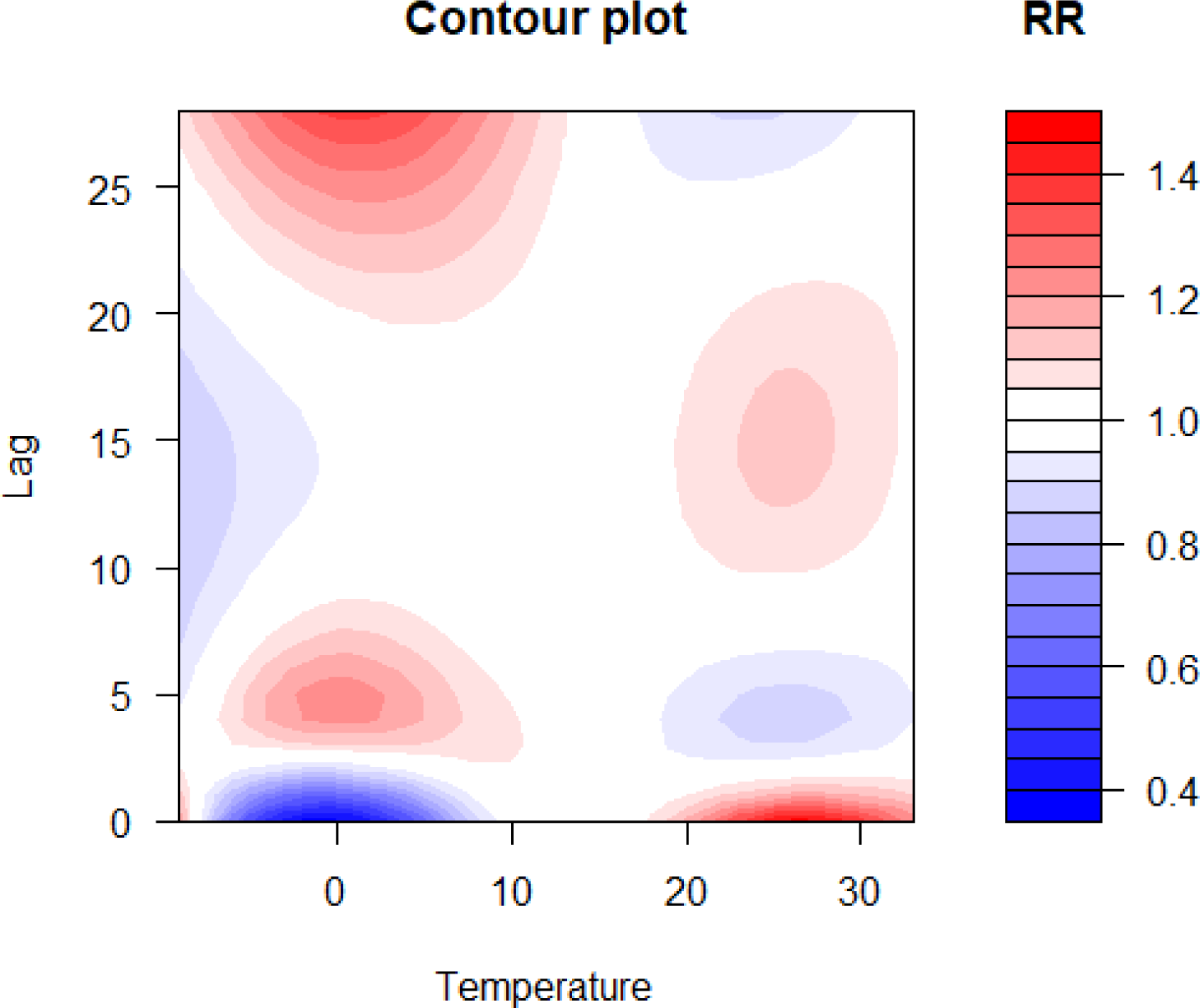
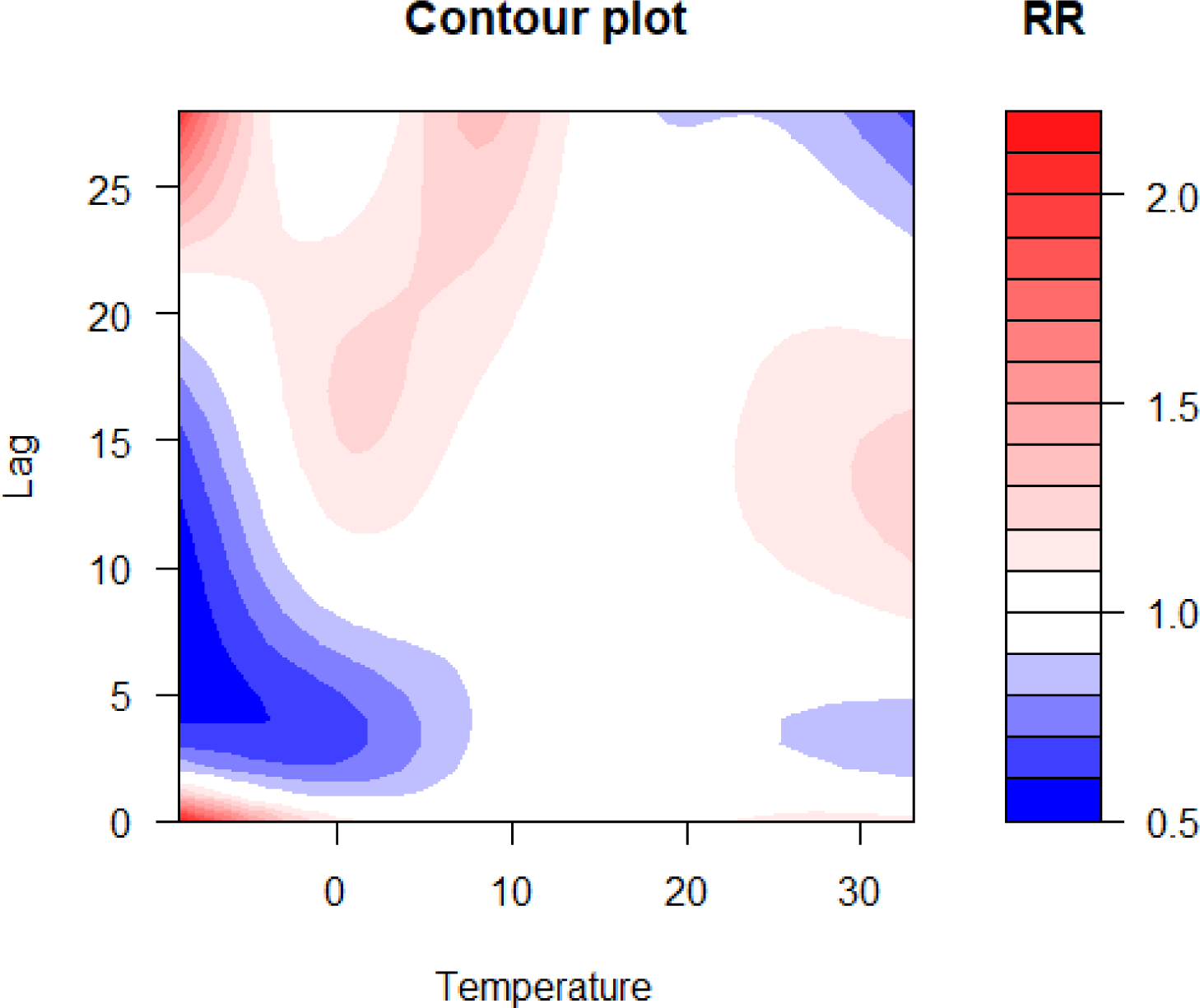
Contour plots of lag-response to temperature based on the age stratification (A: age≤30 group; B: 30<age≤40 group; C:40<age≤50; D:50<age≤60; E: 60<age≤70; F: age>70 group)

**Figure 9.**
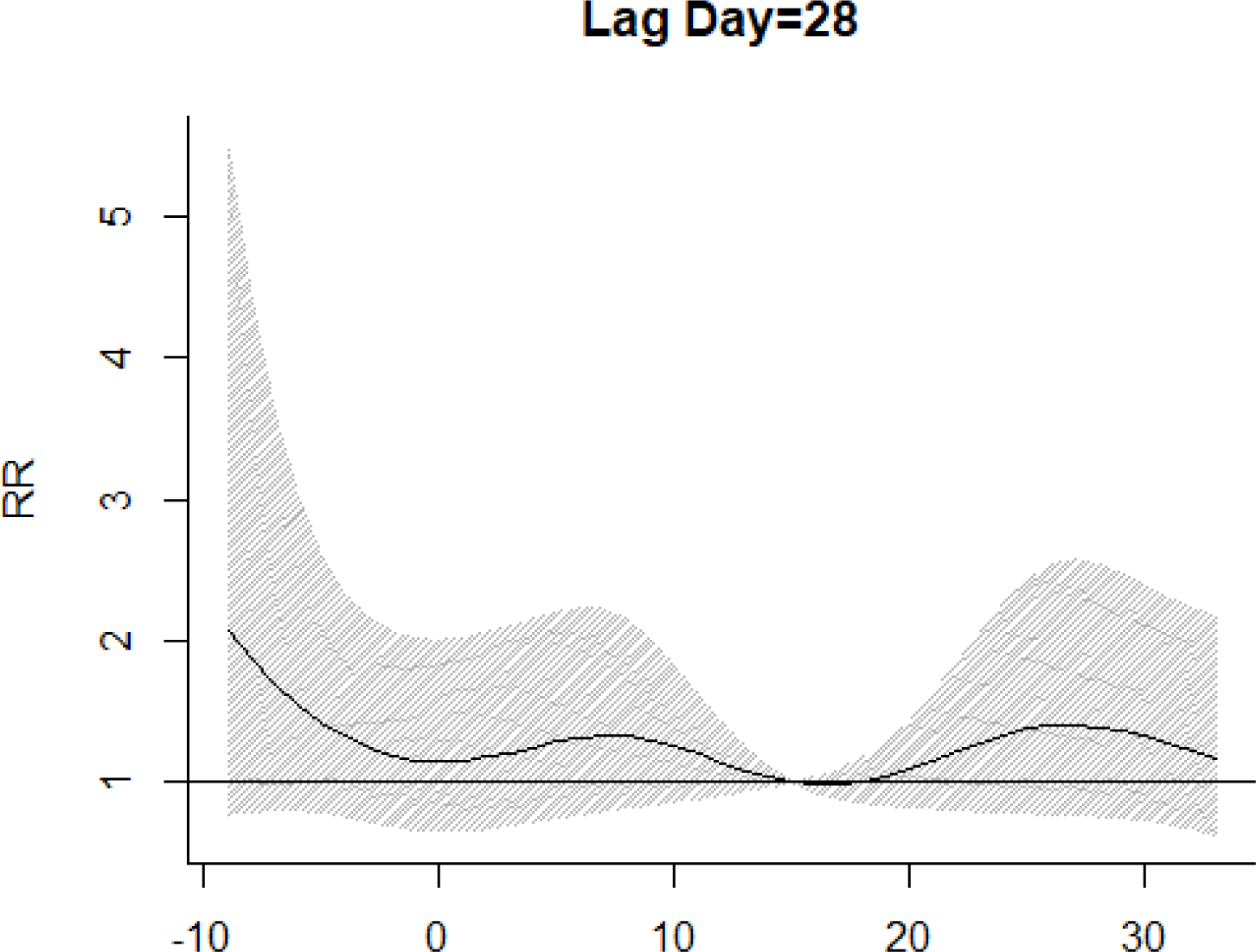
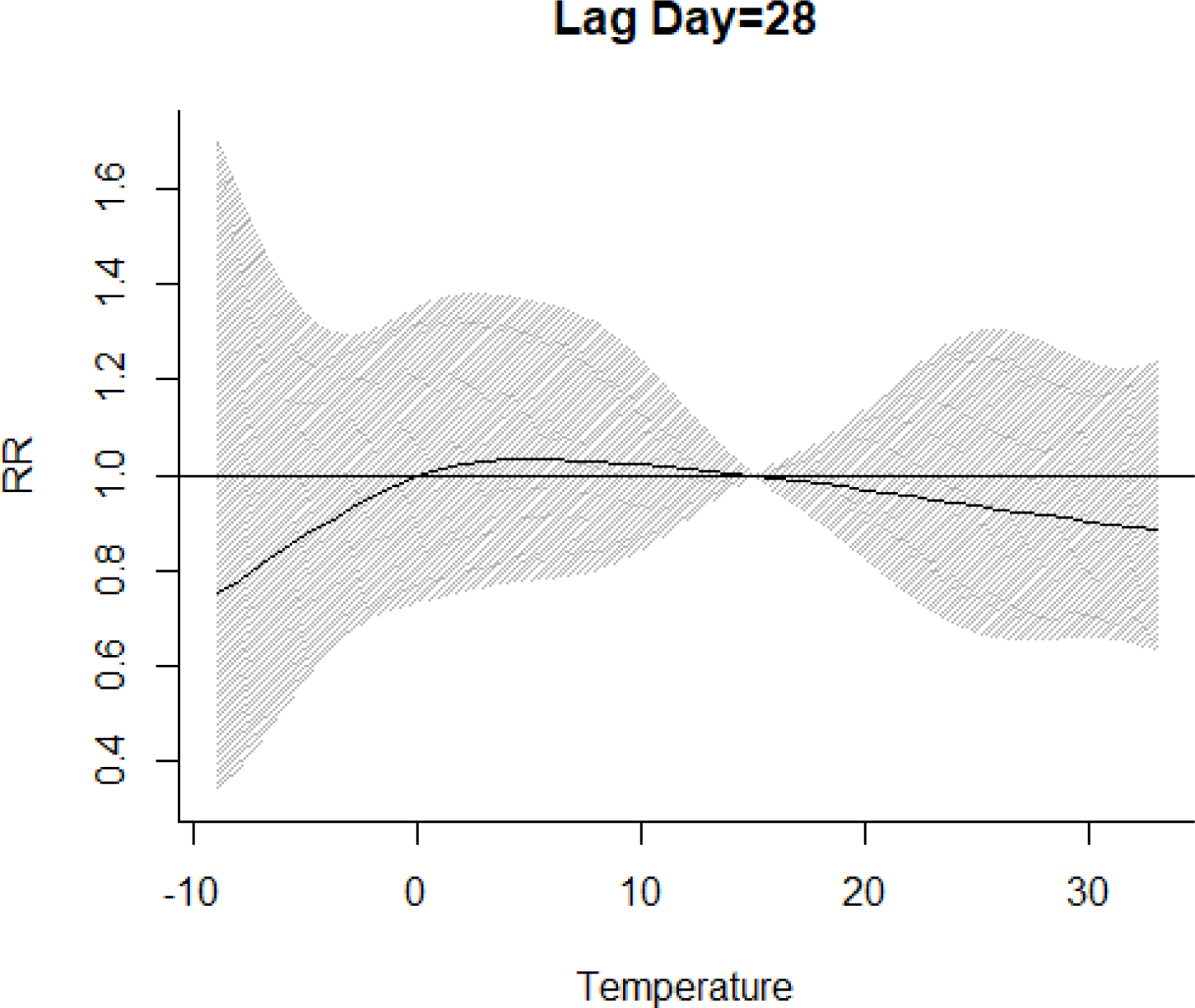
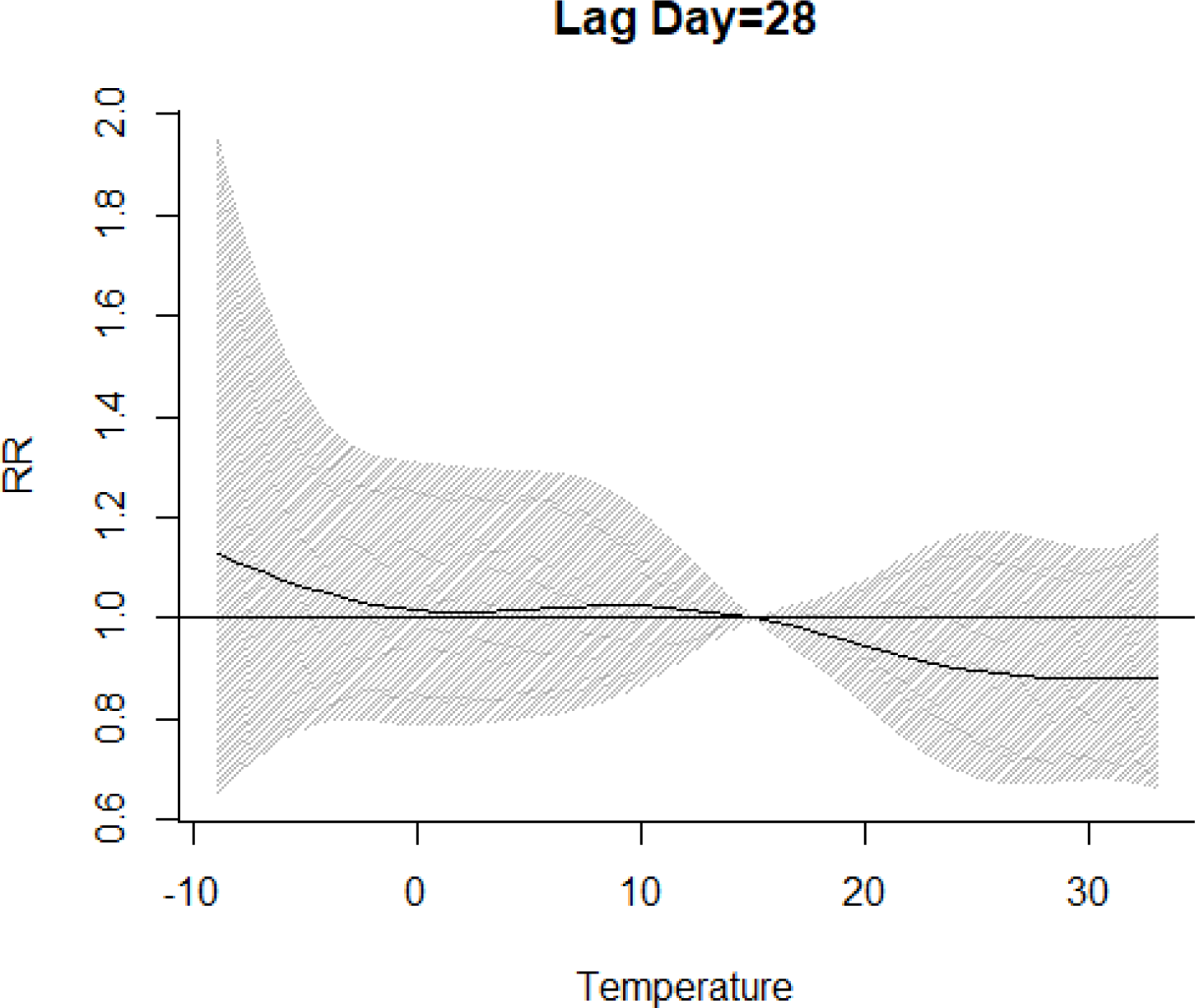
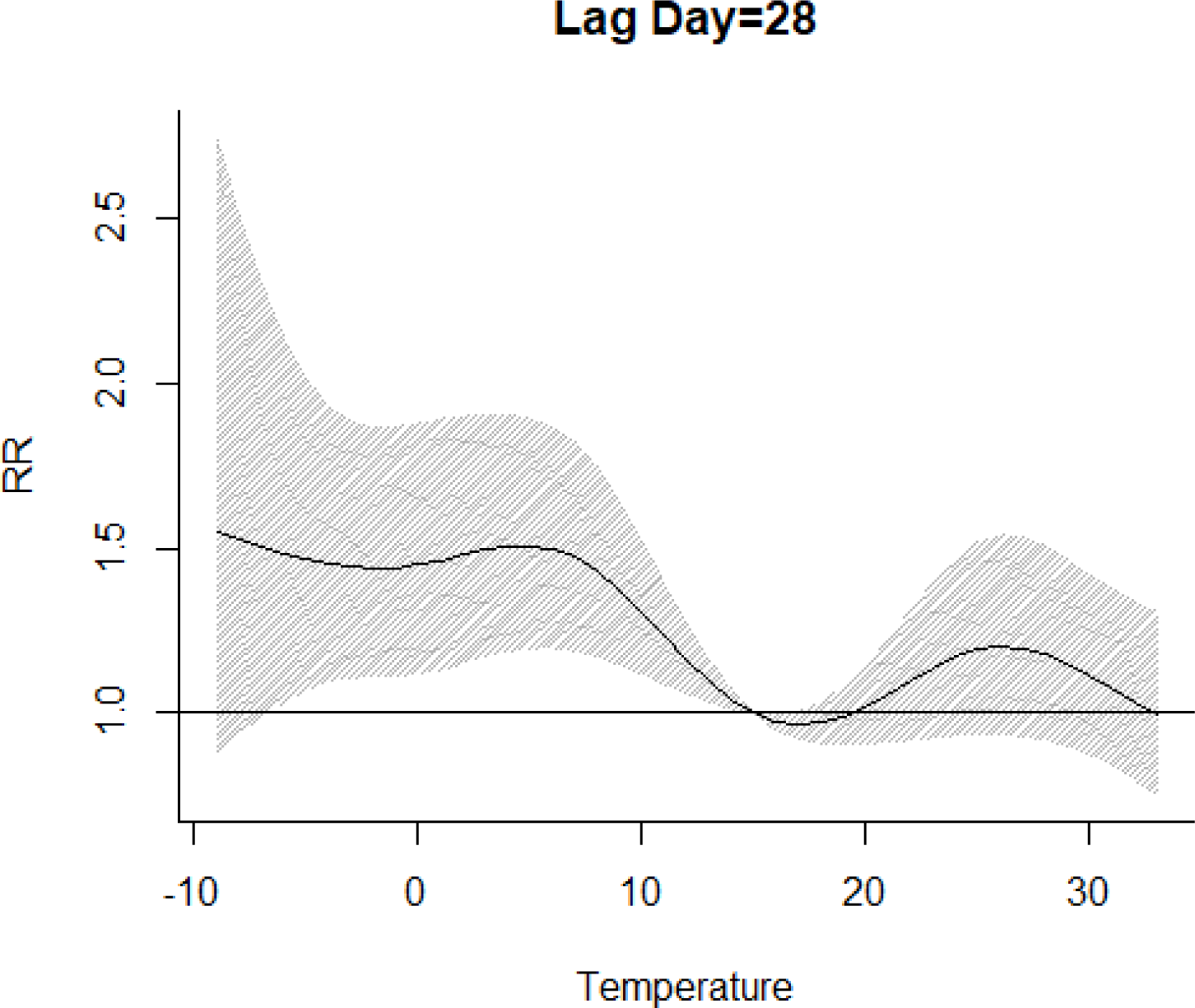
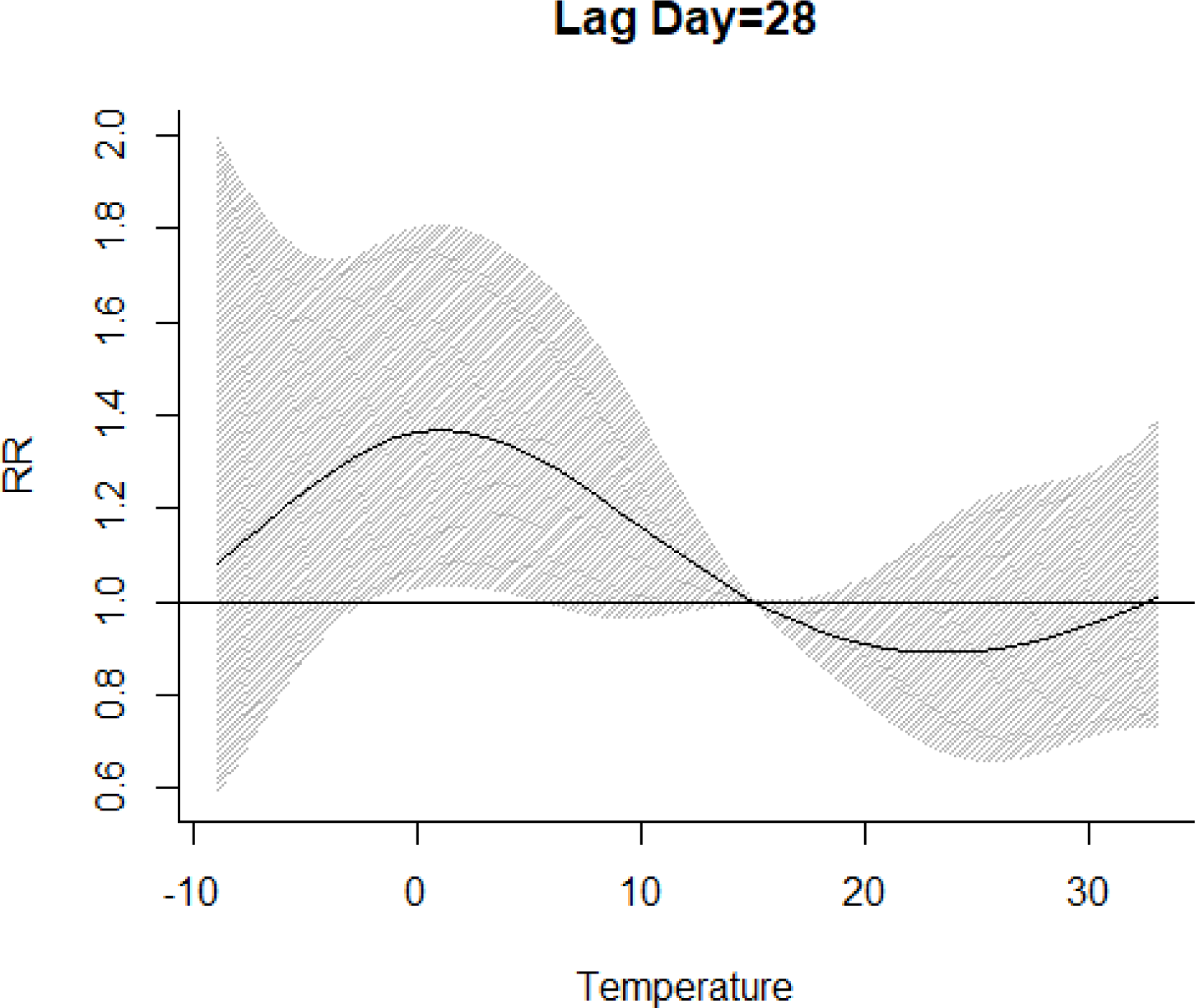
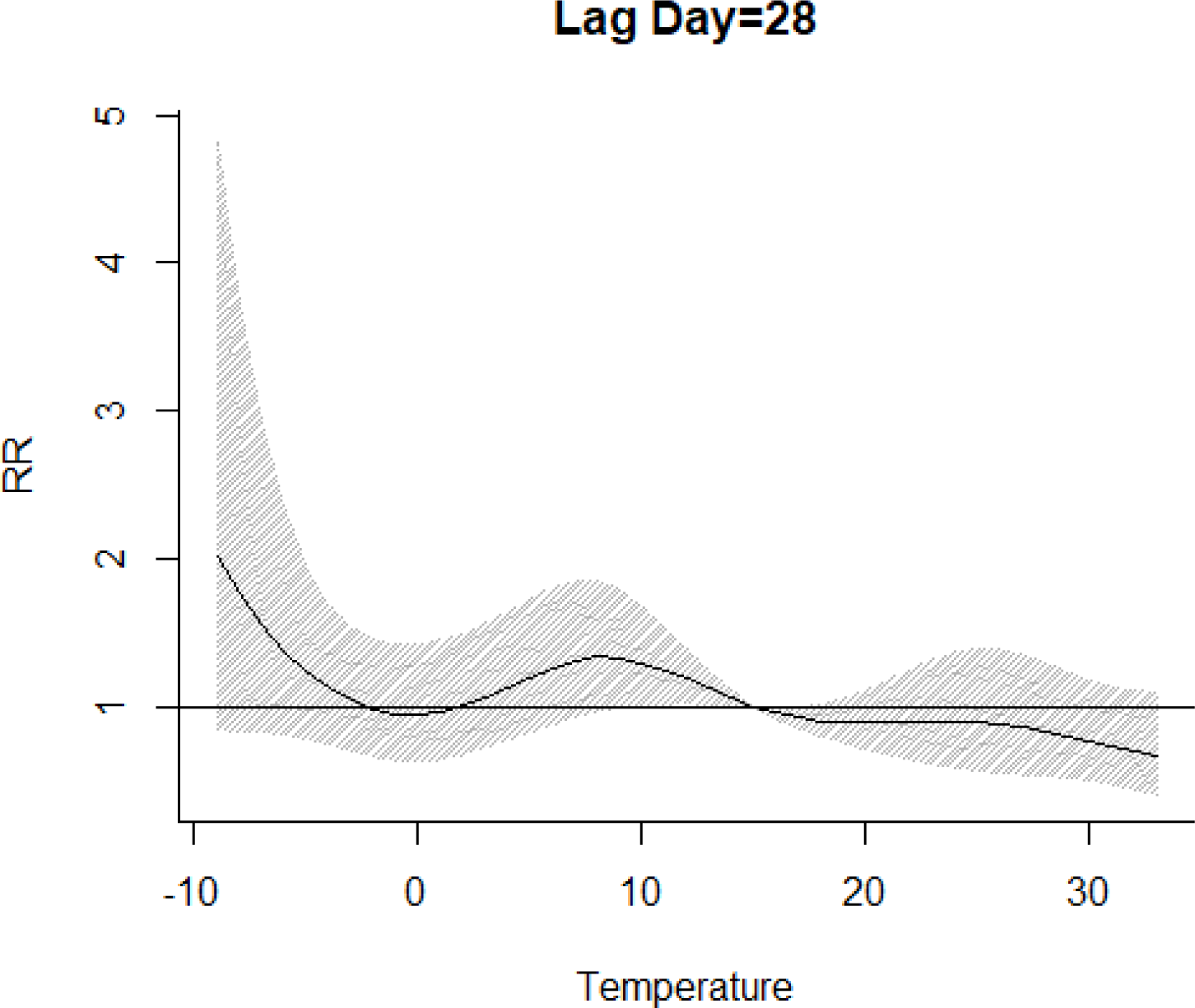
The effect of temperature on the hospitalization when lag day was on 28 (A: age≤30 group; B: 30<age≤40 group; C:40<age≤50; D:50<age≤60; E: 60<age≤70; F: age>70 group)

## Discussion

Lumbar disc herniation (LDH), which the early symptoms are low back pain, has a serious impact on people’s quality of life[5, 20-23]. LDH patients usually take some medicine in their home to relieve their pain, including low back pain, while when intensity of pain was abruptly worsened, they would turn to hospital for relieving pain. In this paper, patients admitted to hospital, were seen that the pain aggravated. The aim of the present research was to examine the relationship between temperature and intensity of back pain among people with lumbar disc herniation (LDH).

In accordance with the present results, previous studies have demonstrated that there was a link between ambient temperature and the sensitivity and intensity of pain, which means that temperature could aggravate back pain[9, 10, 24]. The finding that the proportion of unhealthy habits, drinking and smoking were 5.9% and 7.8%, respectively, which confirmed that previous report that patients with smoking and alcohol abuse had a relative risk of 4.489 and 3.326 for low back pain, respectively, compared with patients without the defined risk factor[25]. Other studies also shown that alcohol use and tobacco use appeared to have a significant relationship with low back pain[23, 26, 27]. We also found that about 16% patients with hypertension, which could provide the evidence that there was the association between hypertension and the prevalence of low back pain[28, 29]. About 6% LBH patients developed the diabetes, which could hint that there might be the link between diabetes and low back pain.[30, 31].

However, some reported previous studies in this field have demonstrated the conflicting results. The reasons which might account for this phenomenon were as follows. Simple correlation was used to evaluate their relationship, but this method reflects the relationship at the same period and neglect the delay effect. On the other hand, some noisy data might influence the result.

The limitation of this study was that we didn’t consider the condition which patients took medicine, as we mentioned above, some LDH patients developed diabetes, hypertension and so on, these could also affect this research. On the other hand, the work category of patients was not considered, we focused on the effect of temperature on low back pain among LDH patients. The future work was to investigate the impact of temperature and relative humidity on low back pain during different seasons.

## Ethical approval

This study was approved by the ethics committee of the Biomedical Engineering Institute, School of Control Science and Engineering, Shandong University. All procedures performed in studies involving human participants were in accordance with the ethical standards of the institutional and/or national research committee and with the 1964 Helsinki declaration and its later amendments or comparable ethical standards.

## Data Availability

Temperature data at the same period was attained from Shandong Environmental Protection Department.

## Acknowledgements

This study was supported in part by grants from the National Natural Science Foundation (#21728701), the Ministry of Education postdoctoral fund (#2015M572044), Shandong Province Science and Technology Development Project (#GG201709260070), the Distinguished Experts of Taishan Scholar Project (grant number ts201511074), China Postdoctoral Science Foundation (grant number 2019M662420), Natural Science Foundation of Shandong Province (grant number ZR2019MH134), Academic Promotion Project of Shandong First Medical University (grant number 2019QL003), Science and Technology Development Program of Shandong Academy of Medical Sciences (grant number 2018-23).

